# Functional respiratory abnormalities in adults who vape daily

**DOI:** 10.1101/2024.10.07.24315006

**Authors:** Ariane Lechasseur, Marc Fortin, Krystelle Godbout, Marie-Ève Boulay, Keven Bergeron, Joanie Routhier, Geneviève Parent-Racine, Annie Roy, Geneviève Boutin, François Maltais, Andréanne Côté, Mathieu C. Morissette

## Abstract

Despite the widespread use of vaping, a very limited number of clinical studies have investigated the effects of this habit on the lungs of healthy individuals. Our group recently initiated the <u>Vap</u>ing <u>A</u>dverse <u>L</u>ung and Heart <u>E</u>vents Coho<u>rt</u> (VapALERT), a prospective study aiming to identify the impacts of vaping on respiratory and cardiovascular health. We elected to report early findings from the pulmonary function tests performed at the initial visit of the first 83 participants recruited so far. Almost 80% of volunteers with no diagnosis of lung disease and who vape daily have an abnormal airway reactivity to metacholine and/or lung clearance index and/or diffusion capacity. We can conclude from this study that adult individuals who vape daily are very likely to present asymptomatic functional respiratory abnormalities, especially airway hyperresponsiveness, ventilation heterogeneity and reduced gas diffusion regardless of past or current tobacco and/or cannabis smoking. Longitudinal studies are crucial to determine how respiratory abnormalities observed in individuals who vape will progress over time.

---

Impacts of vaping on respiratory health are largely unknown. Users can develop “vaping-associated lung illness” (VALI) but this disease remains rare (1). Despite the widespread use of vaping, a very limited number of clinical studies have investigated the effects of this habit on the lungs of healthy individuals. Our group recently initiated the *<u>Vap</u>ing <u>A</u>dverse <u>L</u>ung and Heart <u>E</u>vents Coho<u>rt</u>* (VapALERT), a prospective study aiming to identify the impacts of vaping on respiratory and cardiovascular health. Given the concerning number of respiratory test abnormalities, we elected to report early findings from the pulmonary function tests performed at the initial visit of the first 83 participants recruited so far.

Participants of the VapALERT study are adults who vape daily and who were recruited from the Quebec City metropolitan area. Visits are scheduled every 6 months for a projected 5-year follow-up. Volunteers complete a detailed questionnaire to fully capture their vaping, tobacco and cannabis use as well as their perceived respiratory symptoms. They are subjected to pulmonary function tests at study enrolment including spirometry, lung volumes and diffusion capacity (DLCO) measurements, methacholine challenge test to determine the provocative concentration causing a 20% fall in forced expiratory volume in the first second (PC20), multiple breath nitrogen washout to measure lung clearance index (LCI), fractional exhaled nitric oxide (FeNO) measurement as well as sputum and blood cell counts and differentials.

Mean age of volunteers was 31 years old, 55% being female, with a reported daily vaping of 4.4 years on average (range 3 weeks to 10 years) at enrolment (Table 1). Fourteen participants reported active tobacco smoking (≥7 cigarettes/week), 46 reported being former smokers and 22 never smokers. As for cannabis smoking, 24 reported active use, 28 were past users and 31 never using. Of the 83 participants, 11 had received a previous diagnosis of asthma and 2 of chronic obstructive pulmonary disease (COPD). Six were newly diagnosed with asthma following testing and respirology consultation. Amongst the 64 participants without lung disease diagnosis, 35 (55%) had a PC20 below 16 mg/ml, 25 (39%) had a LCI above or equal to 7.5, and 17 (27%) had a DLCO below 80% of predicted values. Overall, 50 (78.1%) participants without diagnosed lung disease had pulmonary function test abnormalities. As expected, all individuals with newly diagnosed asthma or previously diagnosed asthma or COPD had various abnormalities in measurements of PC20, LCI and DLCO. Six (9%) participants without diagnosed lung disease had blood eosinophils higher or equal to 0.3 × 10^9^/L. Thirty-two (50%) users without diagnosed lung disease were able to produce a valid sputum sample: 3 (9%) had sputum eosinophil levels greater than 2% and 5 (16%) had neutrophils levels greater than 64.4%. Seven (11%) participants without known lung disease had an elevated FeNO (≥25 ppb).

Most participants reported no or mild respiratory symptoms (**Table 1**). A few reported significant elevations in specific symptom scores but there was no association between respiratory symptoms and pulmonary function tests. Spirometry was unremarkable with 63 out of 64 (98%) volunteers with no diagnosed lung disease within normal predicted values (FEV_1_ >80% predicted and FEV_1_/FVC >70% predicted). When using lower limit of normality (LLN) thresholds for FEV_1_/FVC, 58 (91%) individuals remained within normal range.

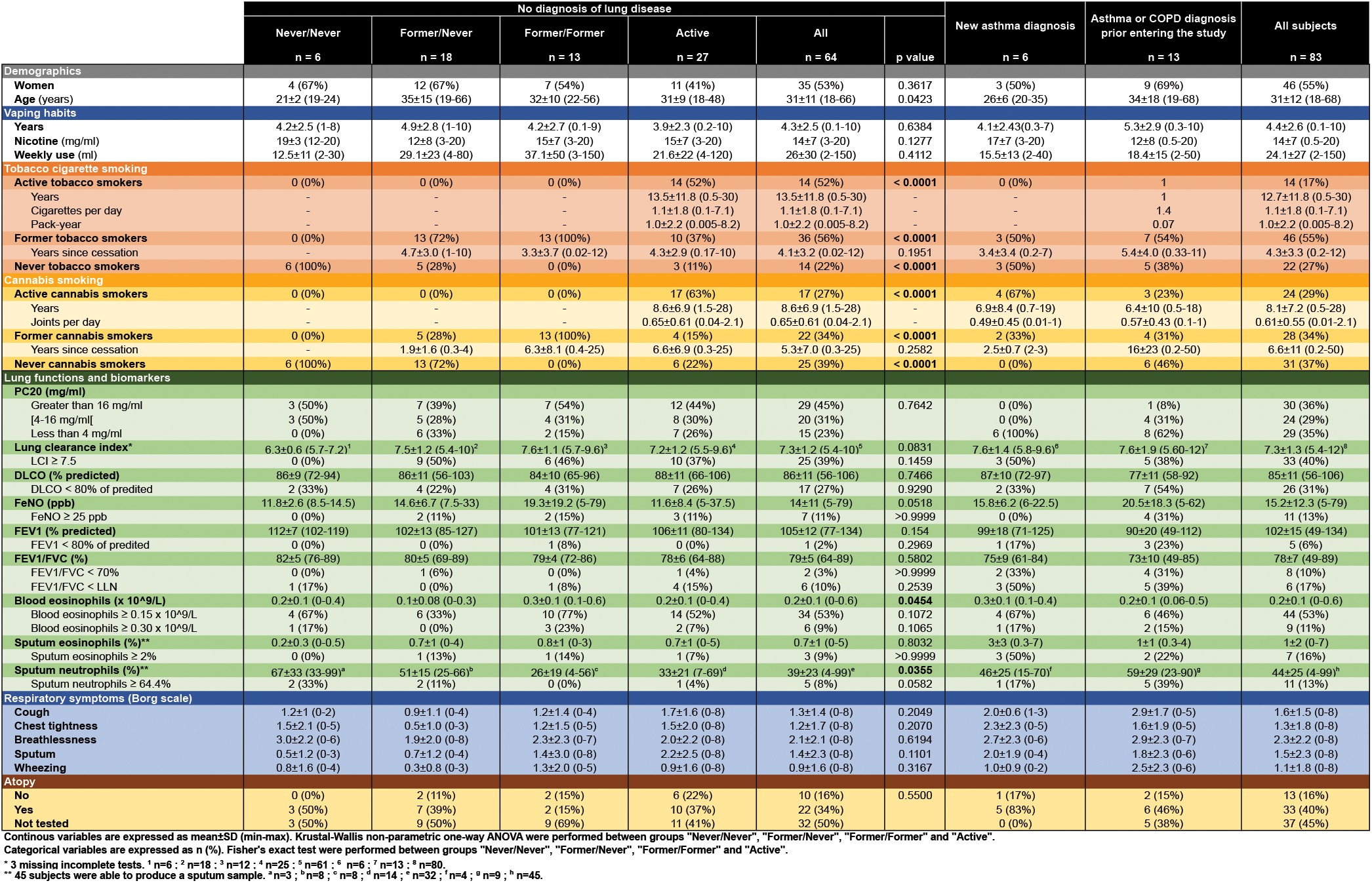

To account for confounding habits of tobacco and cannabis smoking, participants with no diagnosis of lung disease were divided into four groups: Group *“Never/Never*” [never smoked tobacco or cannabis regularly; n=6], Group *“Former/Never*” [stopped smoking tobacco or cannabis and never smoked tobacco or cannabis on a regular basis; n=18], Group *“Former/Former*” [stopped smoking tobacco and cannabis; n=13] and Group *“Active*” [Actively smoking tobacco and/or cannabis; n=27] (**Table 1**). Most adults who vape in our cohort have a history or tobacco and/or cannabis smoking or are actively smoking tobacco and/or cannabis. However, when considering the number of cigarette and/or joint smoked on a daily basis, the great majority have a history of occasional tobacco and recreational cannabis smoking. No significant differences were observed on respiratory tests measurements and proportions of abnormal tests between the four groups, except for lung clearance index where no volunteers of the Never/Never group had abnormal value. However, this could be due to the low number of volunteers in this group. Given vaping is the dominant habit in all volunteers, this supports that history of or current use of tobacco and/or cannabis smoking is unlikely to be driving the observed functional respiratory abnormalities on their own.

This study has no control group with individuals who do not vape, mainly due to the difficulty of properly matching each individual who vape with the proper non-vaping individual while considering all metrics and habits. Instead, volunteers were investigated using established tests with known limits of normality. Since this study recruits from the community, there is a possibility of selection bias towards selecting individuals who are more symptomatic or more concerned that vaping could impact their health. However, considering overall respiratory symptoms are low in the cohort, this possibility is unlikely.

It is definitely concerning that almost 80% of volunteers with no diagnosis of lung disease and who vape daily have an abnormal PC20 and/or LCI and/or DLCO. Positive PC20 denotes airway hyperresponsiveness (AHR) and suggests vaping could affect airway and smooth muscle homeostasis. This is supported by pre-clinical studies that showed exposure to vaping aerosols causes AHR in mice (2, 3). Also, growing evidence supports that vaping could be associated to or interact with asthma (4). Elevated LCI suggests involvement of peripheral airways causing uneven distribution of ventilation and has been associated to several lung diseases including asthma and COPD, and may be more sensitive than spirometry to detect early pulmonary changes caused by smoking (5). A conservative 7.5 upper limit for LCI normality has been used based on studies with healthy volunteers (6, 7). Interestingly, airway hyperresponsiveness and ventilation heterogeneity could be connected since increased ventilation heterogeneity has been associated with worse airway hyperresponsiveness (8). The majority of individuals with low DLCO also had positive PC20 and increased LCI, suggesting reduced gas diffusion could represent a further step of the ongoing process.

There is still a lot to uncover on the effects of vaping on the respiratory system, including interactions with other respiratory habits such as tobacco and cannabis use. However, we can conclude from this study that adult individuals who vape daily are very likely to present asymptomatic functional respiratory abnormalities, especially airway hyperresponsiveness, ventilation heterogeneity and reduced gas diffusion regardless of past or current tobacco and/or cannabis smoking. Longitudinal studies are crucial to determine how respiratory abnormalities observed in individuals who vape will progress over time.

## Data Availability

All data produced in the present work are contained in the manuscript

